# Impact of Anxiety and Depression on Migraine-Related Disability: Results from the Headache Assessment via Digital Platform in the United States (HeAD-US) Study

**DOI:** 10.1101/2025.07.14.25331532

**Authors:** Adalmina Sarkar, Elham Ghanbarian, Kristina Fanning, Alexandre Urani, François Cadiou, Richard B. Lipton, Ali Ezzati

## Abstract

**Background:** Anxiety and depression are common psychiatric comorbidities among people with migraine and may contribute to increased disease burden. However, their impact on both disability during attacks and burden between attacks is not well-characterized in real-world studies. We aimed to assess the relationship between symptoms of anxiety and depression and both migraine-related disability and interictal burden in a large real-world sample of individuals with migraine.

**Methods:** We used cross-sectional data from 6267 participants with migraine recruited using the Migraine Buddy app to create the Headache Assessment via Digital Platform in the United States (HeAD-US) registry. Disability was assessed using the Migraine Disability Assessment Scale (MIDAS), and interictal burden was measured using the Migraine Interictal Burden Scale (MIBS). Anxiety and depressive symptoms were evaluated using the 4-item Personal Health Questionnaire (PHQ-4). Participants were categorized into four subgroups: anxiety symptoms only, depressive symptoms only, both, or neither. Negative binomial regression was used to explore associations between subgroups and migraine disability or burden.

**Results:** Participants were 41.5 ± 13 years old; 90.8% were women. Coexisting anxiety and depression were more common in chronic than episodic migraine (30.3% vs. 20.0%, *p* < 0.001). In adjusted models, anxiety symptoms alone were associated with an 8.7% increase in MIDAS scores and an 11.7% increase in MIBS scores; depressive symptoms alone with 28.4% and 15.5% increases, respectively; and coexisting anxiety and depressive symptoms with 32.5% higher MIDAS scores and 22.6% higher MIBS scores (all *p* < 0.05).

**Conclusions:** In this large real-world study, anxiety and depression were more prevalent in chronic migraine and independently associated with greater migraine-related disability and increased burden between attacks. These findings extend results from population-based and clinic studies to a digital health cohort.

## Introduction

Migraine is a highly prevalent and disabling neurological disorder that affects approximately 12% of the global population and is a leading cause of years lived with disability worldwide (1, 2). Characterized by recurrent headaches and migraine-specific associated symptoms, along with frequent neuropsychiatric comorbidities, migraine significantly impairs quality of life and contributes to substantial personal and societal burden (1).

Anxiety and depression are among the most common psychiatric comorbidities in individuals with migraine, complicating diagnosis, management, and treatment response (3). Anxiety symptoms are reported in up to 47% of migraine patients, with increased risks of generalized anxiety disorder, panic disorder, social anxiety, and phobias (4-6). Meta-analyses confirm that individuals with migraine are significantly more likely to experience anxiety compared to those without (3). Similarly, depression is frequently observed in patients with migraine, with prevalence estimates ranging from 8.6% to 47.9% across studies (7), and multiple investigations have shown elevated rates of major depressive disorder (6, 8-10).

Beyond their high prevalence, anxiety and depression are linked to increased migraine-related disability and overall disease burden (4, 11). In a population study, migraine and depression independently predict decrements in health-related quality of life (12). Rates of psychiatric comorbidities are higher in individuals with chronic migraine (≥15 headache days/month) compared to those with episodic migraine (3, 9). A bidirectional relationship has been demonstrated: migraine is associated with higher rates of incident depression and depression is associated with an elevated rate of incident migraine in longitudinal health plan-based studies (10, 13-16). While some studies propose that migraine-related pain and reduced quality of life contribute to depressive symptoms, others suggest that depression itself may worsen migraine symptoms or increase their frequency (10, 17, 18). A longitudinal population-based study shows that depression is associated with an increased risk of migraine progression from episodic to chronic migraine, and that risk increases with depression severity (18).

Furthermore, anxiety sensitivity—a heightened fear of anxiety-related sensations—has been identified as a marker of migraine burden, associated with both attack severity and interictal impairment (19). Similarly, depression has been independently linked with increased interictal burden, suggesting an effect that extends beyond headache episodes themselves (20).

Given their impact on migraine-related outcomes, and the availability of effective pharmacologic and nonpharmacologic treatments, anxiety and depression are increasingly recognized as modifiable risk factors for migraine onset and progression. Both have been shown to reduce the effectiveness of headache treatments and are associated with worse clinical trajectories (21). Preventing the onset or progression of anxiety disorders may mitigate disability and improve quality of life in people with migraine (13). Moreover, depression has been identified as a predictor of transformation from episodic to chronic migraine, and behavioral interventions targeting mood symptoms can reduce headache frequency and improve outcomes (18, 22).

Despite recognition of anxiety and depression as common comorbidities in migraine, their contribution to both ictal (during-attack) and interictal (between-attack) burden is not well-characterized in real-world populations. The Headache Assessment via Digital Platform in the United States (HeAD-US) registry offers a large cohort of U.S. adults with migraine who track their symptoms using a mobile app. Notably, the cohort includes a much higher proportion of individuals with chronic migraine than typically seen in the general population, and a higher distribution of monthly headaches among those with episodic migraine.

In this study, we used cross-sectional data from 6,267 HeAD-US participants to examine the association between psychiatric symptoms and both headache-related disability and interictal burden, using validated instruments (MIDAS and MIBS). We hypothesized that anxiety and depressive symptoms—particularly when co-occurring—would be associated with higher levels of disability and burden, and that these effects would be more pronounced among individuals with higher monthly headache frequency.

## Methods

### STUDY DESIGN AND PARTICIPANTS

This was a cross-sectional analysis using data from the Headache Assessment via Digital Platform in the United States (HeAD-US) cohort (23). Participants were recruited through the Migraine Buddy smartphone application, which is used to track headache symptoms and management. The study population was limited to adults residing in the United States.Recruitment participants began in September 2023 and lasted for 4 months in total, with users invited to participate via in-app notifications.

Participation in the HeAD-US study was voluntary. Respondents provided electronic informed consent and completed a comprehensive questionnaire within the app. The survey included attention-check questions and predefined response ranges to ensure data quality. Eligible participants were individuals who self-reported a diagnosis of migraine and met diagnostic criteria based on the validated AMS/AMPP diagnostic module, which aligns with the migraine criteria of the International Classification of Headache Disorders, 3rd edition (ICH) (24). All study procedures were approved by the Institutional Review Board (CIRBI Protocol #Pro00072897).

### STATISTICAL ANALYSIS

#### Migraine Frequency and Severity

Headache frequency was based on self-reported average monthly headache days over the previous three months. Participants were categorized as having episodic migraine (EM, <15 days/month) or chronic migraine (CM, ≥15 days/month). Migraine severity was assessed using the Migraine Symptom Severity Scale (MSSS), which evaluates the frequency of seven migraine features. Higher MSSS scores reflect greater symptom burden (9).

#### Migraine Disability and Interictal Burden

Disability was measured using the Migraine Disability Assessment Scale (MIDAS), which sums responses to five items assessing days of missed or reduced productivity over the past three months (9). MIDAS scores were grouped into four grades: I (0–5), II (6–10), III (11–20), and IV (≥21), indicating increasing levels of disability (25). Interictal burden was measured using the 4-item Migraine Interictal Burden Scale (MIBS), assessing functional limitations between attacks. Responses are weighted by severity, and total scores are categorized as low (0–2), moderate (3–4), or severe (≥5) burden (26).

#### Depression and Anxiety

Psychiatric symptoms were assessed using the 4-item Patient Health Questionnaire (PHQ-4), which includes two items for anxiety and two for depression (27). A subscore ≥3 on the anxiety items defined as presence of anxiety symptoms; similarly, a subscore ≥3 on the depression items defined as presence of depressive symptoms (28). Participants were grouped into four categories: symptoms of neither anxiety nor depression, anxiety symptoms only, depressive symptoms only, or both.

#### Other Covariates

Additional covariates included demographic characteristics and clinical features. Age was treated as a continuous variable. Sex was categorized as *male, female*, or *prefer not to answer*. Race was categorized as *White or Caucasian only, Asian only, Black or African American only, American Indian or Alaska Native, Other – two or more races*, and *Prefer not to answer*. Annual household income was reported in increments and categorized as *<$25,000, $25,000–$50,000, $50,000–$75,000, $75,000*–*$100,000*, or*>$100,000*, and *Prefer not to answer*. Allodynia was defined based on self-reported pain or unpleasant skin sensations during headaches occurring at least ‘some of the time’ or more frequently, using a single-item question with response options: ‘none,’ ‘rarely,’ ‘some of the time,’ and ‘half the time or more’ (9). Additional covariates included the Migraine Symptom Severity Score (MSSS) and monthly headache-day frequency, both treated as continuous variables.

#### Statistical Analysis

All analyses were conducted using IBM SPSS Statistics version 25.0 (IBM Corp., Armonk, NY). Two-tailed hypothesis testing was used throughout, and statistical significance was defined as *p* < 0.05. Missing data was minimal and did not warrant imputation. Descriptive statistics were calculated for all variables, with means and standard deviations reported for continuous variables (age, MIDAS, MIBS, MSSS) and frequencies with percentages reported for categorical variables (sex, income, race, allodynia, MIDAS grades, MIBS grades, anxiety symptoms, depressive symptoms).

Independent-sample *t*-tests and chi-square tests were used to compare demographic and clinical characteristics between participants with episodic migraine (EM) and chronic migraine (CM). Comparisons across psychiatric subgroups (neither, anxiety symptoms only, depressive symptoms only, both) were conducted using one-way analysis of variance (ANOVA) for continuous variables and chi-square tests for categorical variables, followed by post-hoc pairwise comparisons where appropriate.

Negative binomial regression models were used to examine associations between psychiatric subgroup and MIDAS and MIBS scores due to overdispersion in the data.Covariates in the adjusted models included age (continuous), sex (male/female/prefer not to answer) race (White vs nonwhite), income (<$75,000 vs. ≥$75,000), cutaneous allodynia (present vs. absent, defined as experiencing pain or unpleasant skin sensations during headaches at least “some of the time”), Migraine Symptom Severity Score (MSSS; continuous), and monthly headache-day frequency (continuous, 1–30 days/month).

To visualize the association between psychiatric symptoms and migraine-related disability and interictal burden, predicted MIDAS and MIBS scores were calculated from the fully adjusted negative binomial regression models. These predictions were generated across the full range of monthly headache days (1–30) for each psychiatric symptom subgroup, using the model coefficients. Covariates such as age, sex, income, race, allodynia, and MSSS were held constant at their mean or reference values, allowing visualization of the independent effect of psychiatric symptoms and headache frequency on predicted scores. The resulting graphs illustrate the additive relationship between psychiatric symptom burden and headache frequency on disability (MIDAS) and interictal burden (MIBS).

#### Bias and Study Size

As a digital platform study relying on self-reported data, this analysis is subject to potential selection and reporting biases. Individuals using a migraine tracking app may differ systematically from the general population. Psychiatric symptoms and migraine burden were measured using validated but brief screening tools (PHQ-4, MIDAS, MIBS), which may not capture the full clinical spectrum. The sample was a convenience sample of eligible respondents enrolled over a defined time period; no formal sample size calculations were performed.

## Results

### Baseline Demographics

The study sample included 6,267 participants with a mean age of 41.5 years (SD = 13.0); 90.8% identified as women and 84.6% as white. The mean monthly headache frequency was 11.7 days (SD = 8.2). Based on monthly headache frequency, 62.6% were classified as having episodic migraine (EM) and 37.4% as having chronic migraine (CM).

Most participants reported severe migraine-related disability (MIDAS grade IV: 79.8%) and severe interictal burden (MIBS grade IV: 74.8%). Cutaneous allodynia was reported by 53.5% of the sample, and the mean Migraine Symptom Severity Score (MSSS) was 18.1 (SD = 2.2).

Regarding psychiatric symptoms, 29.9% of participants screened positive for depressive symptoms, and 42.5% for anxiety symptoms. Distribution by psychiatric subgroup was neither anxiety nor depressive symptoms (51.4%), anxiety symptoms only (18.7%), depressive symptoms only (6.1%), and both anxiety symptoms and depressive symptoms (23.8%).

### Comparison Between EM and CM Groups

Compared to participants with EM, those with CM had significantly more headache days (20.8 vs. 6.3 per month, *p* < 0.001), a greater proportion with severe disability (MIDAS grade IV; 94.8% vs. 70.9%, *p* < 0.001) and severe interictal burden (MIBS grade IV; 86.1% vs. 68.0%, *p* < 0.001), and a greater relative frequency of cutaneous allodynia (64.0% vs. 47.2%, *p* < 0.001).CM participants were also more likely to report both anxiety and depressive symptoms (30.3% vs. 20.0%, *p* < 0.001) and depressive symptoms only (8.0% vs. 4.9%, *p* < 0.001). There was no significant difference in anxiety symptoms alone between groups. MSSS did not significantly differ by migraine subtype (Table 2).

**Table 1.** Baseline Demographics and Headache Characteristics

| Characteristic | Respondents (N = 6,267) § |
| --- | --- |
| Age, years, mean (SD) | 41.54 (13.067) |
| Women, n (%) | 5692 (90.8) |
| Race, n (%) |  |
| White or Caucasian only | 5299 (84.6) |
| Black or African American only | 184 (2.9) |
| Asian only | 117 (1.9) |
| American Indian or Alaska Native only | 37 (0.6) |
| Other - two or more races | 392 (6.3) |
| Prefer not to answer | 238 (3.8) |
| Annual household income, n (%) |  |
| <\$25,000 | 603 (9.6) |
| \$25,000-50,000 | 945 (15.1) |
| \$50,000-75,000 | 908 (14.5) |
| \$75,000-100,000 | 899 (14.3) |
| >\$100,000 | 1629 (26.0) |
| Prefer not to answer | 1283 (20.5) |
| Cutaneous allodynia present, ¶ n (%) | 3351 (53.5) |
| MSSS, ¶ mean (SD) | 18.09 (2.220) |
| MIDAS Category |  |
| MIDAS score grade I/II, n (%) | 530 (8.5) |
| MIDAS score grade III, n (%) | 730 (11.6) |
| MIDAS score grade IV, n (%) | 5003 (79.8) |
| MIBS Category |  |
| MIBS score grade I/II, n (%) | 785 (12.5) |
| MIBS score grade III, n (%) | 795 (12.7) |
| MIBS score grade IV, n (%) | 4687 (74.8) |
| Monthly headache day frequency |  |
| Days/month, mean (SD) | 11.68 (8.217) |
| Category, n (%) |  |
| 0-3 days/month | 1005 (16.0) |
| 4-7 days/month | 1492 (23.8) |
| 8-14 days/month | 1427 (22.8) |
| ≥15 days/month | 2343 (37.4) |
| †Depressive symptoms, n (%) | 1874 (29.9) |
| ‡Anxiety symptoms, n (%) | 2665 (42.5) |
| Psychiatric subgroups, n (%) |  |
| Neither anxiety nor depressive symptoms | 3220 (51.4) |
| Anxiety symptoms only | 1173 (18.7) |
| Depressive symptoms only | 381 (6.1) |
| Anxiety and depressive symptoms | 1492 (23.8) |

†Depressive symptoms were defined as a subscore ≥3 on depressive items from the 4-item Personal Health Questionnaire (PHQ-4).
‡Anxiety symptoms were defined as a subscore ≥3 on anxiety items from the PHQ-4.
¶ Allodynia was defined as experiencing pain or unpleasant skin sensations during headaches at least some of the time; MSSS = Migraine Symptom Severity Score; MIDAS = Migraine Disability Assessment Scale (continuous scores from 0 to 270, grades I-IV); MIDAS grades are defined as follows: Grade I/II (little to mild disability, scores 0-10), Grade III (moderate disability, 11-20), and Grade IV (severe disability, scores ≥21); MIBS = Migraine Interictal Burden Scale (continuous scores from 0 to 12, grade I-IV); MIBS grades are defined as follows: Grade I/II (no or mild interictal burden, scores 0-2), Grade III (moderate interictal burden, scores 3-4), and Grade IV (severe interictal burden, scores 5-12)

**Table 2.**
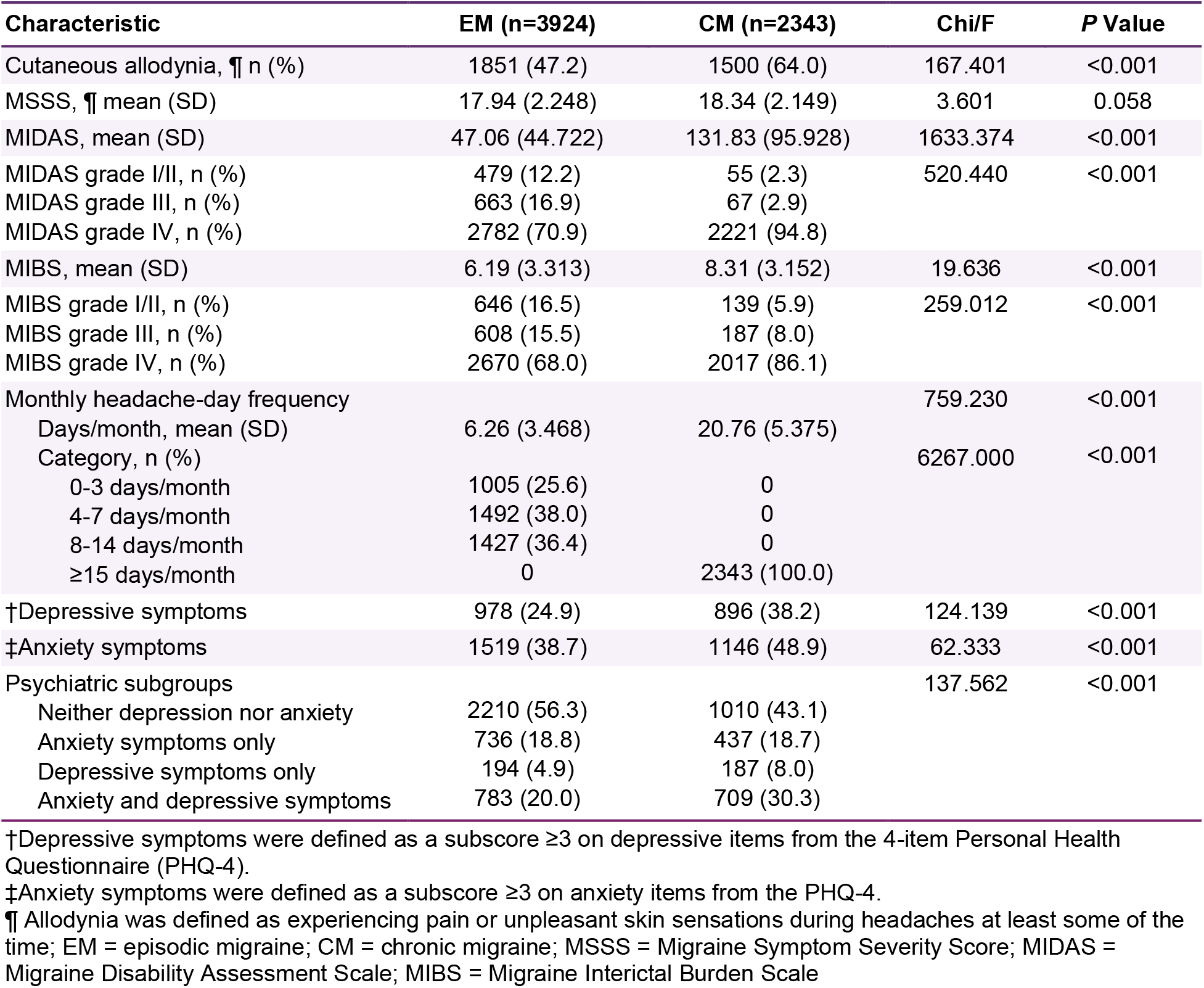
Baseline Headache and Psychiatric Characteristics by Episodic and Chronic Migraine

### Comparison by Psychiatric Symptom Subgroups

There were significant demographic and clinical differences across psychiatric subgroups. Participants with both anxiety symptoms and depressive symptoms were younger (mean age 37.9 years) and had lower household income compared to those with neither symptom. The relative frequency of allodynia was lowest in the group with no comorbidity, intermediate in the groups with depressive or anxiety symptoms and highest in group with both comorbidities (*p* < 0.001). MSSS increased in the groups with comorbidity. Mean monthly headache frequency was lowest in the group without symptoms of depression or anxiety and was highest in the groups with depression alone or with both symptoms (*p* < 0.001).

Disability and interictal burden also increased with psychiatric comorbidity. Mean MIDAS scores were 64.5 in those with neither symptom, 73.7 in those with anxiety symptoms only, 105.6 in those with depressive symptoms only, and 106.6 in those with both (*p* < 0.001). Mean MIBS scores followed a similar pattern (6.2, 7.2, 7.8, and 8.3 respectively, *p* < 0.001). Participants with both symptoms were more likely to have grade IV disability and burden. (Table 3)

**Table 3.** Baseline Demographics and Headache Characteristics by Psychiatric Subgroup

| Parameter | Characteristic | Neither anxiety<br>nor depressive<br>symptoms<br>n=3220 (51.4) | Anxiety<br>symptoms only<br>n=1173 (18.7) | Depressive<br>symptoms only<br>n=381 (6.1) | Anxiety &<br>depressive<br>symptoms<br>n=1492 (23.8) | Chi/F | P- Value |
| --- | --- | --- | --- | --- | --- | --- | --- |
| Sex | Male | 269 (8.4) | 67 (5.7) | 36 (9.4) | 141 (9.5) | 36.817 | <0.001 |
|  | Female | 2933 (91.1) | 1094 (93.3) | 343 (90.0) | 1321 (88.5) |  |  |
|  | Prefer not to answer | 18 (0.6) | 12 (1.0) | 2 (0.5) | 30 (2.0) |  |  |
| Age | Mean (SD) | 44.53 (12.824) | 38.01 (12.113) | 41.28 (13.161) | 37.91 (12.673) | F=130.546 | <0.001 |
| Race | White or Caucasian only | 2753 (85.5) | 985 (84.0) | 310 (81.4) | 1251 (83.8) | 29.339 | 0.015 |
|  | Black or African American only | 86 (2.7) | 30 (2.6) | 15 (3.9) | 53 (3.6) |  |  |
|  | Asian only | 57 (1.8) | 19 (1.6) | 16 (4.2) | 25 (1.7) |  |  |
|  | Native American or Alaska Native only | 16 (0.5) | 12 (1.0) | 2 (0.5) | 7 (0.5) |  |  |
|  | Other - two or more races | 192 (6.0) | 73 (6.2) | 20 (5.2) | 107 (7.2) |  |  |
|  | Prefer not to answer | 116 (3.6) | 54 (4.6) | 18 (4.7) | 49 (3.3) |  |  |
| Annual income | <\$25,000 | 214 (6.6) | 105 (9.0) | 33 (8.7) | 251 (16.8) | 191.235 | <0.001 |
| | \$25,000-50,000 | 386 (12.0) | 179 (15.3) | 87 (22.8) | 293 (19.6) | | |
| | \$50,000-75,000 | 463 (14.4) | 155 (13.2) | 59 (15.5) | 231 (15.5) | | |
| | \$75,000 -100,000 | 497 (15.4) | 190 (16.2) | 48 (12.6) | 164 (11.0) | | |
| | >\$100,000 | 986 (30.6) | 313 (26.7) | 78 (20.5) | 252 (16.9) | | |
|  | Prefer not to answer | 674 (20.9) | 231 (19.7) | 76 (19.9) | 301 (20.2) |  |  |
| Allodynia ¶ | No | 1612 (50.1) | 546 (46.5) | 168 (44.1) | 589 (39.5) | 46.886 | <0.001 |
|  | Yes | 1608 (49.9) | 627 (53.5) | 213 (55.9) | 903 (60.5) |  |  |
| Monthly headache days | Mean (SD) | 10.49 (7.769) | 11.60 (8.043) | 14.00 (8.819) | 13.71 (8.614) | F=64.890 | <0.001 |
| MSSS ¶¶ | Mean (SD) | 17.87 (2.294) | 18.19 (2.138) | 18.23 (2.144) | 18.46 (2.080) | F=25.510 | <0.001 |
| MIDAS | Mean (SD) | 64.52 (71.028) | 73.73 (70.596) | 105.63 (91.306) | 106.58 (91.832) | F=116.594 | <0.001 |
| MIDAS score grade | MIDAS grade I/II | 376 (11.7) | 79 (6.7) | 18 (4.7) | 61 (4.1) | 141.756 | <0.001 |
|  | MIDAS grade III | 435 (13.5) | 146 (12.4) | 34 (8.9) | 114 (7.6) |  |  |
|  | MIDAS grade IV | 2409 (74.8) | 948 (80.8) | 329 (86.4) | 1317 (88.3) |  |  |
| MIBS | Mean (SD) | 6.20 (3.332) | 7.20 (3.303) | 7.75 (3.379) | 8.30 (3.192) | F=147.535 | <0.001 |
| MIBS score grade | MIBS grade I/II | 528 (16.4) | 127 (10.8) | 34 (8.9) | 96 (6.4) | 187.366 | <0.001 |
|  | MIBS grade III | 503 (15.6) | 135 (11.5) | 38 (10.0) | 119 (8.0) |  |  |
|  | MIBS grade IV | 2189 (68.0) | 911 (77.7) | 309 (81.1) | 1277 (85.6) |  |  |

Values are n (%) unless noted otherwise.
†Depressive symptoms were defined as a subscore ≥3 on depressive items from the 4-item Personal Health Questionnaire (PHQ-4).
‡Anxiety symptoms were defined as a subscore ≥3 on anxiety items from the PHQ-4.
¶ Allodynia was defined as experiencing pain or unpleasant skin sensations during headaches at least some of the time; MSSS = Migraine Symptom Severity Score; MIDAS = Migraine Disability Assessment Scale; MIBS = Migraine Interictal Burden Scale

**Table 4.**
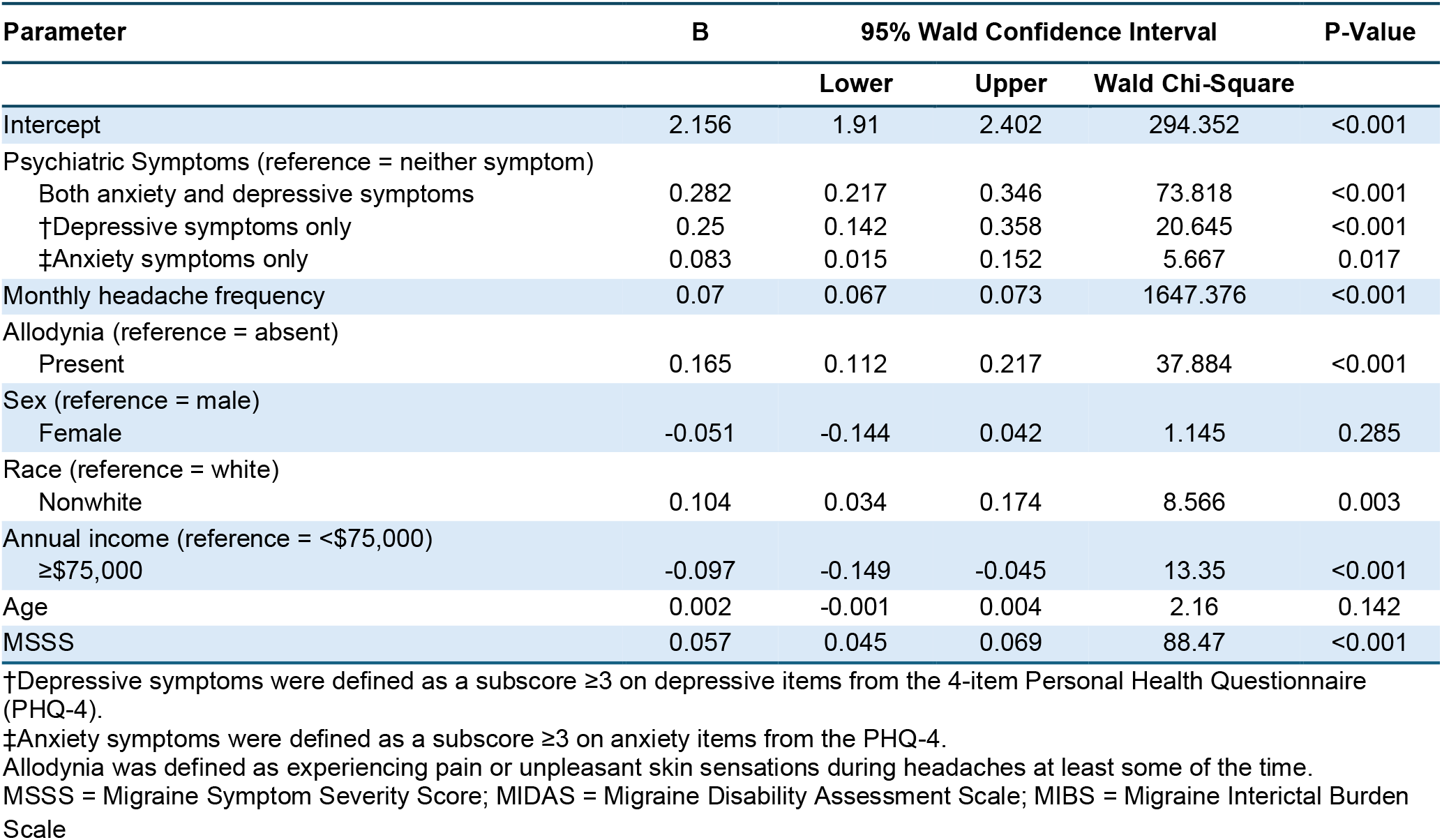
Parameter Estimates for MIDAS Score

**Table 5.** Parameter Estimates for MIBS Score

| Parameter | B | 95% Wald Confidence Interval |  |  | P-Value |
| --- | --- | --- | --- | --- | --- |
|  |  | Lower | Upper | Wald Chi-Square |  |
| Intercept | 0.821 | 0.56 | 1.081 | 38.146 | <0.001 |
| Psychiatric Symptoms (reference = neither symptom) |  |  |  |  |  |
| Both anxiety and depressive symptoms | 0.203 | 0.135 | 0.272 | 33.996 | <0.001 |
| †Depressive symptoms only | 0.144 | 0.030 | 0.258 | 6.093 | 0.014 |
| ‡Anxiety symptoms only | 0.11 | 0.037 | 0.183 | 8.751 | 0.003 |
| Monthly headache frequency | 0.016 | 0.013 | 0.020 | 87.835 | <0.001 |
| Allodynia (reference = absent) |  |  |  |  |  |
| Present | 0.153 | 0.097 | 0.208 | 28.955 | <0.001 |
| Sex (reference = male) |  |  |  |  |  |
| Female | -0.005 | -0.104 | 0.094 | 0.01 | 0.922 |
| Race (reference = white) |  |  |  |  |  |
| Nonwhite | 0.04 | 0.034 | 0.114 | 1.139 | 0.286 |
| Annual income (reference = <\$75,000) | | | | | |
| ≥\$75,000 | -0.038 | -0.093 | 0.017 | 1.8 | 0.18 |
| Age | -0.001 | -0.003 | 0.002 | 0.337 | 0.562 |
| MSSS | 0.044 | 0.031 | 0.056 | 46.774 | <0.001 |

†Depressive symptoms were defined as a subscore ≥3 on depressive items from the 4-item Personal Health Questionnaire (PHQ-4).
‡Anxiety symptoms were defined as a subscore ≥3 on anxiety items from the PHQ-4.
Allodynia was defined as experiencing pain or unpleasant skin sensations during headaches at least some of the time.
MSSS = Migraine Symptom Severity Score; MIDAS = Migraine Disability Assessment Scale; MIBS = Migraine Interictal Burden Scale

### Association Between Psychiatric Symptoms and Migraine-Related Disability

Negative binomial regression adjusting for demographics, headache frequency, MSSS, and allodynia showed that anxiety symptoms alone were associated with an 8.7% increase in MIDAS score (RR=1.087, *p* = 0.017), depressive symptoms alone with a 28.4% increase (RR=1.284, *p* < 0.001), and coexisting symptoms with a 32.5% increase (RR=1.325, *p* < 0.001), compared to those with neither symptom. Additional covariates independently associated with higher MIDAS scores included higher MSSS, presence of allodynia, and lower income levels, indicating that both clinical and social factors contribute to greater migraine-related disability. Predicted MIDAS scores increased in a dose-response fashion across psychiatric subgroups and headache frequency strata (Figure 1a), highlighting the additive burden of psychiatric comorbidities on disability. The group with depressive symptoms only did not differ significantly from the comorbid group (*p* = 0.584). All other pairwise comparisons between psychiatric subgroups yielded statistically significant differences (*p* ≤ 0.005).

**Figure 1a.**
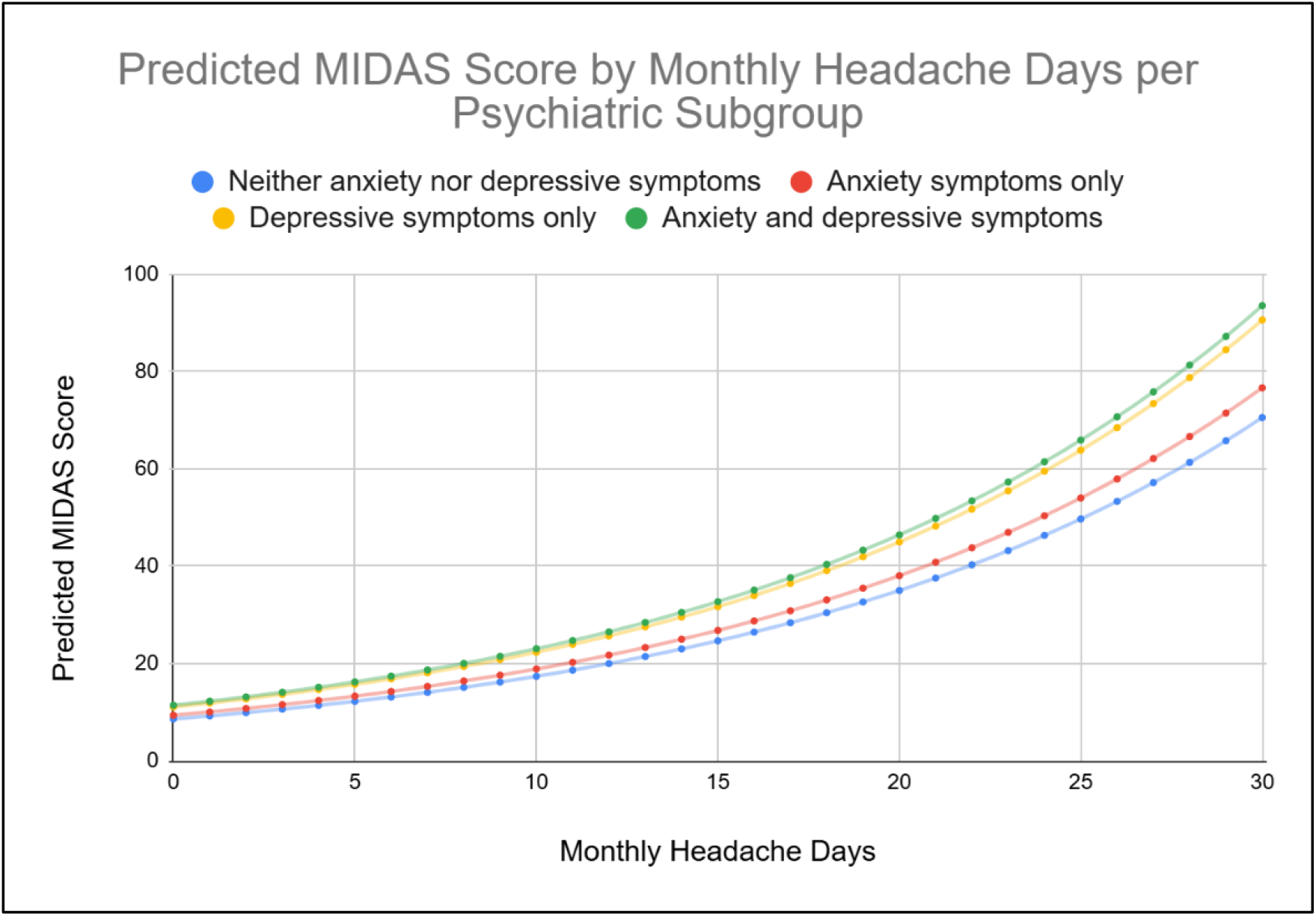
Predicted MIDAS Score by Monthly Headache Day Frequency and Psychiatric Comorbidity Subgroup.

**Figure 1b.**
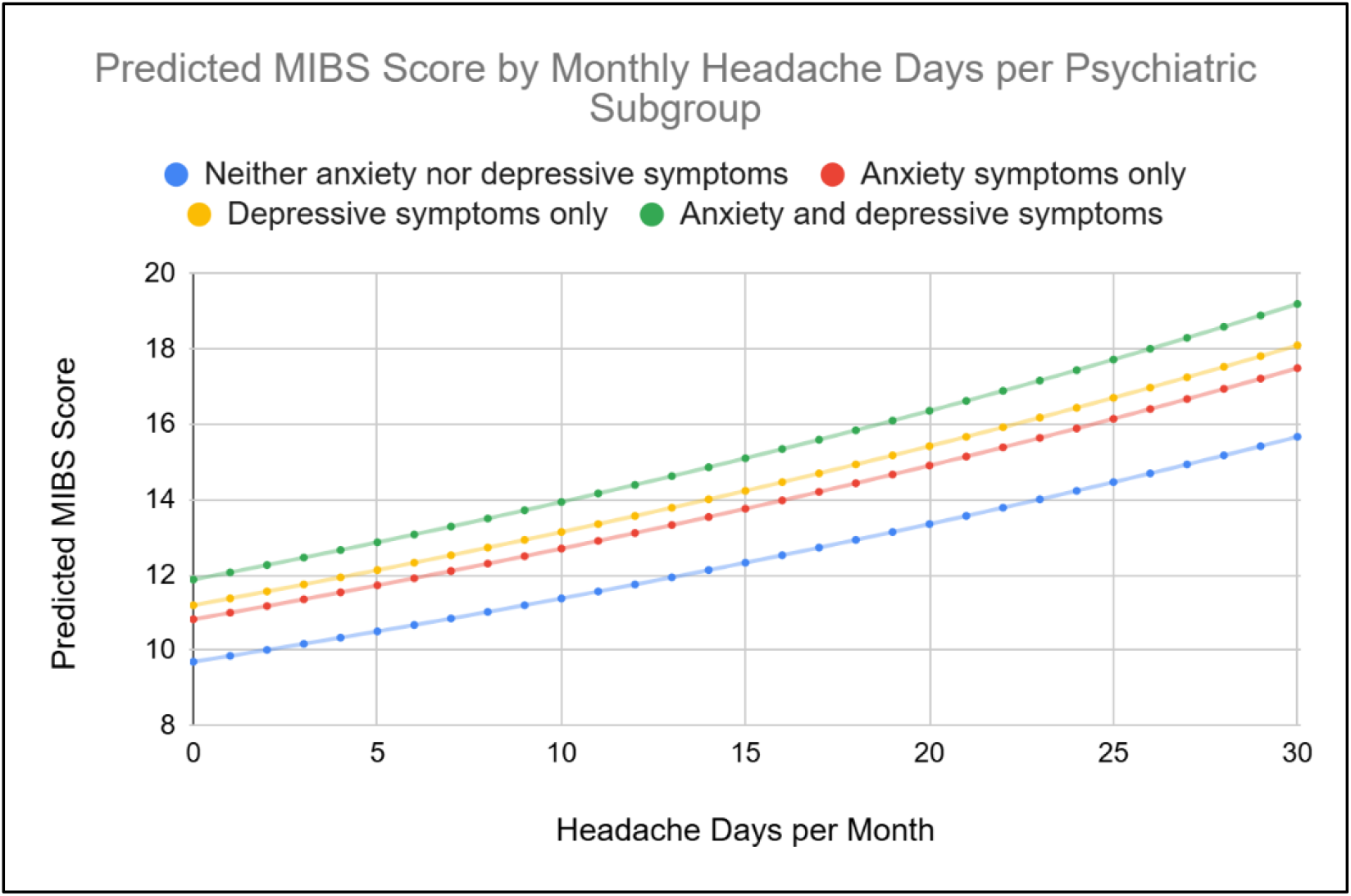
Predicted MIBS Score by Monthly Headache Days per Psychiatric Subgroup.

### Association Between Psychiatric Symptoms and Interictal Burden

In adjusted models, anxiety symptoms alone were associated with an 11.7% increase in MIBS scores (RR=1.117, *p* = 0.003), depressive symptoms alone with a 15.5% increase (RR=1.155, *p* = 0.014), and coexisting symptoms with a 22.6% increase (RR=1.226, *p* < 0.001), relative to participants without either condition. Predicted MIBS scores showed a similar additive effect of psychiatric symptoms, increasing steadily with headache burden and psychiatric symptom load (Figure 5). Allodynia and higher MSSS were also associated with increased interictal burden in the fully adjusted model (Figure 1b), reinforcing the multifactorial contributors to between-attack impact in migraine patients.Pairwise comparisons revealed that the depressive symptom only group did not differ significantly from the comorbid group (*p* = 0.330) or the anxiety symptom only group (*p* = 0.597) but did differ significantly from the group with neither symptom (*p* = 0.014).Additionally, the anxiety symptom only group differed significantly from both the comorbid group (*p* = 0.026) and neither symptom group (*p* = 0.003).

## Discussion

In this large, real-world sample of individuals with migraine, symptoms of anxiety and depressive symptoms were more prevalent among those with chronic migraine compared to those with episodic migraine. Both anxiety symptoms and depressive symptoms were independently associated with greater headache-related disability, with the strongest associations observed in individuals reporting co-occurring symptoms. Notably, depressive symptoms showed a stronger association with disability than anxiety symptoms when examined separately.

Consistent with previous studies, we found that psychiatric symptoms were significantly more prevalent among individuals with chronic migraine (CM) than those with episodic migraine (EM). This pattern echoes findings from the MAST study, which showed that increased headache frequency was linked to higher rates of anxiety, depressive symptoms, and migraine-related disability (3). In our data, depressive symptoms—whether occurring alone or with anxiety—were more strongly associated with CM than anxiety alone, which showed no significant variation by migraine subtype. These results suggest a closer connection between depressive symptoms and CM, though the cross-sectional design limits conclusions about directionality or causation.

The CAMEO study, a large internet-based survey of over 16,000 individuals with migraine, reported a somewhat different pattern—namely, that anxiety symptoms alone were more common in EM than in CM (9). This contrast may be due in part to differences in sample characteristics. Notably, our cohort had a higher proportion of participants reporting severe disability and interictal burden (MIDAS and MIBS grade III or IV), which likely reflects a population with more complex or treatment-refractory migraine. This overrepresentation of severe cases may have attenuated the observed differences in psychiatric symptom burden between EM and CM, particularly in the subgroup with anxiety symptoms alone. We also found that participants with CM were substantially more likely to report moderate to severe levels of migraine-related disability and interictal burden. Both MIDAS scores and MIBS scores increased as MHDs increased though the MIDAS relationship is curvilinear and the MIBS relationship is linear. The shift from lower levels of disability (grade I/II) to high levels of disability (grade III/IV) scores across MIDAS and MIBS underscores the compounding effect of frequent headache days. As the burden of disease increases, individuals may experience reduced autonomy and functional impairment, potentially contributing to the emergence or worsening of depressive symptoms over time (10, 20). This trend mirrors results from the CAMEO study, which also reported that CM respondents were more likely to fall into the higher disability categories.

Across our sample, anxiety symptoms were more commonly reported than depressive symptoms, regardless of migraine subtype—a trend that aligns with several prior studies. For instance, the FRAMIG study, a population-based survey from France, found that 28% of respondents reported anxiety symptoms alone, compared to just 3% with depression alone (4). In contrast, the CAMEO study observed the opposite pattern, with a higher proportion of individuals reporting depression alone (9.6%) than anxiety alone (7.2%) (9). These discrepancies across studies may reflect differences in population characteristics, recruitment methods, or cultural perceptions of psychiatric symptoms.

Both anxiety and depressive symptoms were independently associated with increased migraine-related disability, with the strongest effect observed among individuals reporting both symptoms. This finding aligns with prior research underscoring the additive burden of psychiatric comorbidities in migraine. Interestingly, although anxiety symptoms were more prevalent in our sample, depressive symptoms—either alone or in combination with anxiety— had a stronger association with disability. This pattern is further reflected in our regression models: depressive symptoms alone were linked to a 28.4% increase in MIDAS scores, while co-occurring anxiety and depression were associated with a 32.5% increase; in contrast, anxiety symptoms alone were associated with only an 8.7% increase compared to the neither symptoms group (*p* < 0.05). Pairwise comparisons of MIDAS scores showed that the depressive symptom only group did not differ significantly from the comorbid group, while all other comparisons between psychiatric subgroups were statistically significant. This pattern suggests that depressive symptoms may account for a substantial portion of the migraine-related disability observed in individuals with both anxiety and depressive symptoms. Our findings align with prior studies showing that anxiety and depression are linked to increased migraine-related disability. The CAMEO study reported a similar pattern, with the highest disability among those with symptoms of both anxiety and depression, followed by depression alone, then anxiety alone (9). However, effect sizes were larger in CAMEO—co-occurring symptoms were associated with a 79% increase in disability, compared to 32.5% in our study. This difference may reflect the higher baseline disability in our sample, where nearly 80% had MIDAS grade IV, possibly limiting the observable impact of psychiatric symptoms. In addition, the use of PHQ-4 provides a less robust assessment of anxiety symptoms than measures used in some earlier studies (29). The MAST study also found elevated rates of anxiety and depression among people with migraine compared to those without (3). In contrast, our analysis focused solely on individuals with migraine, assessing variation in disability across psychiatric symptom subgroups. This narrower scope may explain the smaller relative effect sizes observed in our data.

A key observation from our study was the increase in both migraine-related disability and interictal burden with rising headache frequency and psychiatric symptom severity. Rather than increasing at a constant rate, predicted MIDAS scores rose more sharply as monthly headache days and symptom burden increased, reflecting the compounding impact of psychiatric symptoms and headache frequency. This pattern is consistent with the log-link function used in negative binomial regression and the larger coefficient for headache frequency in the MIDAS model, which results in steeper predicted score increases. These findings underscore the potential value of integrated interventions that jointly target headache frequency and psychiatric symptoms to mitigate overall migraine burden (18, 22). In contrast, predicted MIBS scores increased in a more linear fashion. This may partly reflect the structure of the MIBS questionnaire, which measures burden between attacks; at high headache frequencies, the number of attack-free days—and thus the scope for interictal burden—diminishes. As such, the relationship between headache frequency and MIBS may plateau at higher frequencies, limiting further additive increases.

Pairwise comparisons across psychiatric subgroups revealed a more nuanced picture for interictal burden. The depressive symptom only group did not differ significantly from either the comorbid group or the anxiety symptoms only group but did differ from the group with neither symptom. Additionally, the anxiety symptoms only group differed significantly from both the comorbid and the neither symptom group. These findings suggest that while comorbid symptoms are associated with the highest levels of interictal burden, the distinct contributions of anxiety and depressive symptoms may be more difficult to disentangle. This may reflect the nature of interictal burden itself, which captures affective and anticipatory symptoms between attacks, where anxiety may play a larger or more persistent role than during the attacks themselves (20, 30). However, the presence of depressive symptoms appears to contribute additively to burden when co-occurring with anxiety, as evidenced by the significant difference between the comorbid and anxiety-only groups. This is consistent with a previous study that found the interictal period was uniquely associated with depression, possibly due to secondary factors of migraine such as having to give up plans and social activities (20).

Several limitations should be noted. First, all data were self-reported, which may introduce recall or reporting bias. Second, psychiatric symptoms were assessed using the PHQ-4, a brief screening tool that may not capture the full spectrum or severity of anxiety and depression. In addition, psychiatric symptoms may be influenced by medications respondents are taking. Third, the cross-sectional design limits our ability to infer causality or temporal relationships between psychiatric symptoms and migraine-related outcomes. Finally, the study sample included a relatively high proportion of individuals with CM, which may overestimate overall disability levels while potentially underestimating the relative impact of psychiatric symptoms.

## Conclusion

This study has important implications for both clinical care and public health. Although the American Headache Society and other clinical guidelines acknowledge the frequent co-occurrence of anxiety and depression in individuals with migraine, there remains no standardized approach to screening or managing these comorbidities in routine care. Current treatment strategies are variable, and the absence of integrated guidelines may contribute to inconsistent care and increased healthcare costs (31). Our findings highlight the strong association between psychiatric symptoms and both migraine-related disability and interictal burden, emphasizing the need for treatment models that jointly target mood and migraine symptoms. Future research should aim to clarify the temporal and potentially causal relationship between psychiatric comorbidities and migraine outcomes.

Longitudinal studies that track changes in mood symptoms and headache burden over time, along with measures such as anxiety sensitivity, may offer deeper insight into mechanisms and inform early intervention strategies.

## Data Availability

All data produced in the present study are available upon reasonable request to the authors

